# Prevalence of Neutralizing Antibodies against Adenoviruses types -C5, -D26 and -B35 used in vaccination platforms, in Healthy and HIV-Infected Adults and Children from Burkina Faso and Chad

**DOI:** 10.1101/2022.06.07.22276076

**Authors:** Aline Raissa Ouoba, Océane Paris, Chatté Adawaye, Guy Takoudjou Dzomo, Abderrazzack Adoum Fouda, Dramane Kania, Amidou Diarra, Zenaba Abdramane Kallo, Isidore Tiandiogo Traore, Sodiomon Bienvenu Sirima, Edouard Tuaillon, Philippe Van de Perre, Eric J. Kremer, Franck Jean Daniel Mennechet

## Abstract

Vaccines derived from human adenoviruses (HAdV) are currently being used and trialed against numerous infectious agents. However, pre-existing humoral immunity can impair vaccines efficacy and safety. Strategies to circumvent this immunity often involve the use of vectors with lower seroprevalence. We evaluated HAdV-C5, HAdV-D26 and HAdV-B35 seroprevalence from healthy and HIV-infected populations from Burkina Faso and Chad. Seroprevalence for HAdV-C5 was high and comparable between countries (54%-66%), and the highest in the HIV-infected groups from sub-Saharan regions (∼90%). However, compared to France (4%), seroprevalence for HAdV-D26 was significantly higher in sub-Saharan groups (∼47%). By contrast, HAdV-B35 seroprevalence was low for all groups tested. We also found that HAdV-D26 complexed with immunoglobulins induced strong activation of dendritic cells in vitro. Our study fills gaps in the epidemiological data needed to optimize HAdV-derived vaccines in sub-Saharan regions, and highlights the necessity to better adjust vaccination strategies in Africa.

**Article summary line:** HAdVs Seroprevalence in Burkina Faso and Chad

## Introduction

Vaccines save millions of lives every year and limit major financial and public health system losses (1). As of April 4^th^ 2022, ∼17% (25/153) of COVID-19 vaccines are viral vector-derived (2). Among them, five are human adenovirus (HAdV) based, including Convidecia (CanSino Biologics), Sputnik V (Gamaleya Research Institute), and the Johnson & Johnson/Janssen (3). The most widely distributed COVID-19 vaccine is Vaszevria (ChAdOx1-S), a chimpanzee adenovirus-derived vaccine developed by Oxford University/AstraZeneca. HAdVs were among the first vectors developed for gene transfer/vaccines. Among the ∼100 ∼HAdV types, HAdV species C type 5 (HAdV-C5) is the best characterized and is the most widely used (4).

A drawback in the use of HAdV-derived vaccines is pre-existing immunity that impairs vaccine efficacy and potential immunotoxicity (5,6). Current data suggest that both innate and adaptive immune responses impact the duration of transgene expression from HAdVs (7). Natural infection or the administration of AdV vectors in pre-immune subject results in an activation of innate immune responses associated with the production of pro-inflammatory cytokines/chemokines, following by an adaptive immunity, including T and B cell responses. Although HAdV-induced T cells are responsible for the lysis of cells expressing viral and transgenic products (8), anti-HAdV Abs favor capsid uptake by antigen-presenting cells through Fc receptors (9,10). HAdV-C5-based vaccines have therefore shown an inverse correlation between efficacy/safety and pre-existing humoral immunity. A better understanding of the interaction between innate and pre-existing immunity, vaccines, is a major objective in the development of Ad-derived vaccines. Several strategies have been deployed to bypass humoral immunity, including genetic and chemical modifications of the capsid, generation of chimeric vectors, and the use of AdV with low seroprevalence (6). Alternative candidates to HAdV-C5 are numerous, but only HAdV-D26, -B35 and -E4 are currently in clinical trials/use (11). HAdV-D26-based vaccines have been extensively tested in multiple clinical programs (12,13). Depending on the trial, HAdV-D26-derived vaccines are generally used in prime-boost heterologous strategies with the modified vaccinia Ankara or other HAdV types of such as HAdV-B35 or HAdV-C5. For COVID-19 vaccine, Ad26.COV2.S is a single dose (13), and Sputnik is a heterologous vaccine using HAdV-D26 with -C5 in a prime-boost regimen (14). HAdV- B35 on other hand is not among candidates of COVID-19 vaccines, but has long been part of the vaccination landscape for many infectious diseases, in particularly HIV (15), Ebola virus (16), malaria (17) and tuberculosis (18), which are particularly devastating in sub-Saharan Africa.

While HAdV-C5 seroprevalence is well documented, data on HAdV-D26 and HAdV-B35 are less extensive (11). For HAdV-D26, accumulating evidence shows that seroprevalence in Europe and North America rarely exceeds 10% (11,19). The few studies conducted on the seroprevalence of HAdV-D26 in Africa have shown seroprevalence between 10 and 90%, with low or intermediate NAb titers compared to HAdV-C5 (20–22). For HAdV-B35, seroprevalence is generally low worldwide, with rates slightly higher in some sub-Saharan regions (11,23,24). However, global mapping of the seroprevalence of HAdV-D26 and HAdV-B35 remains incomplete. These gaps make HAdV-derived COVID-19 vaccines less predictable in populations that are globally more affected by infectious diseases.

In this study, we quantified HAdV-C5, -D26 and -B35 seroprevalence in individuals from Burkina Faso and Chad. In contrast to Europe & North America, our study indicates that HAdV-D26 seroprevalence is high, particularly in people living with HIV. By contrast, HAdV-B35 seroprevalence is low in all the cohorts tested. Moreover, immunoglobulin- complexed HAdV-D26s stimulate the activation of human phagocytes.

## Methods

### Sample location and management

This cross-sectional study was conducted on 424 plasma samples from Burkina Faso (241), Chad (93) & France (50) **(Table 01)**. Samples were collected from adults or children, HIV- infected, or HIV-uninfected donors. Samples from HIV-uninfected subjects in Burkina Faso were collected in 2012 at the National Malaria Research and Training Center (CNRFP) and the Centre MURAZ in 2017. Collection and use of plasma samples for this study was approved by institutional review boards of the CNRFP (AEP-04/07/20 1O/CIB-CNRFP) and Institutional Ethic Committee of the Centre MURAZ (N°2017-361/MS/SG/CM/DG). For Chad, HIV-uninfected samples were collected at the “Bon Samaritain” hospital (from December 2018 to November 2019). In France, samples were provided by the French Blood service (EFS, France) and the study was approved by the Occitanie & Midi-Pyrenees EFS scientific board (EFS-OCPM: N°21PLER2019-0002). Samples from HIV-infected subjects from Burkina Faso were randomly selected from the ANRS 12174 Promise-PEP trial sample collection (20), in accordance with the sub-study request established and approved by Institutional Ethic Committee of the country. For Chad, samples from HIV-infected subjects were collected in 2019 at the Sectorial AIDS Control Program Center (PSLS) in the city of N’Djamena. All samples were supported by a written and signed informed consent.

**Table 01:** Demographics data

| **Regional Sample** | **<u>Overall</u>** | **<u>Burkina Faso</u>**<br>**Bobo-dioulasso**<br>**/Ouagadougou** | **<u>Chad</u>**<br>**N'Djamena** | **<u>France</u>**<br>**Montpellier** |
| --- | --- | --- | --- | --- |
| **Number of Sample (%)** | 424 (100) | 281 (66,3) | 93 (21,9) | 50 (11,8) |
| **Sex, n (% , %)** |  |  |  |  |
| Male | 154 (36,3) | 107 (30,1) | 35 (37,6) | 12 (36) |
| Female | 252 (59,5) | 174 (69,9) | 58 (62,4) | 20 (26) |
| unknown | 18 (4,2) | 0 | 0 | 18 (38) |
| **Age range,**<br>**Mean Age** | 7 months - 82 years<br>28,6 years | 10 months - 82 years<br>27,9 years | 7 months - 59 years<br>30,2 years | 20 - 55 years<br>29,5 years |
| **Age, year; n (%)** |  |  |  |  |
| (0-9) | 48 | 37 (13,2) | 11 (11,8) | 0 |
| (10-17) | 51 | 49 (17,4) | 2 (2,2) | 0 |
| (18-25) | 100 | 62 (22,1) | 21 (22,6) | 17 (34) |
| (26-44) | 131 | 78 (27,7) | 43 (46,2) | 10 (20) |
| (≥45) | 75 | 55 (19,6) | 15 (16,1) | 5 (10) |
| Unknown | 19 | 0 | 1 (1,1) | 18 (36) |
| **HIV status, n (%)** |  |  |  |  |
| Negative | 358 (84,4) | 261 (92,8) | 47 (50,5) | 50 (100) |
| Positive | 66 (15,6) | 20 (7,2) | 46 (49,5) | 0 (0) |

### Adenoviruses, cells, and culture conditions

For neutralization assays, 911 cells were cultured as described previously (25). For *ex vivo* MoDC stimulation assay, blood was obtained from healthy adult donors via the EFS, Montpellier, France and processed as previously described (26). GFP-expressing HadVs were amplified in (HEK) 293 or 911 cells and purified, and quantified as previously described (27) Vector purity typically reached *>*99%. HAdV-D26-GFP was provided by Dr. Eric Weaver (University of Nebraska-Lincoln, USA) and HAdV-B35-GFP by Andre Lieber (University of Washington, Seattle, USA).

### Adenovirus neutralization assays

For neutralizing antibody (NAb) titration tests, 911 cells were seeded at 10^5^ cells/well in 24-well plates and left to attach for 1 h. Plasma were diluted from 1:40 to 1:2160 and pre-incubated during 30 mins at 20°C with GFP-expressing-HAdV at 15, 100 or 500 pp/cell for HAdV-C5, B35 and D26, respectively. As the seroprevalence of HAdV-B35 is typically low, a pre-screening at 1:10, 1:100 and 1:1000 sample dilutions was performed to select samples of interest. Two-fold dilutions of the HAdV of interest were systematically included in duplicate to control the infectivity capacity of the vectors. HAdV standards, controls, and samples were cultured with cells for 20 h at 37°C and 5% of CO_2_. The next day, the cells were collected and assayed for GFP expression by flow cytometry (FACScalibur®- BD Bioscience or NovoCyte ACEA®- Agilent). The NAb titers was quantified by identifying the sample dilutions that generate 50% of GFP+ cells. Samples were considered positive when NAb triter ≥ 100. Data analyses were performed using the FlowJo software (BD Bioscience, France).

### Immune complexes formation and DC stimulation

Monocyte-derived DCs (MoDCs) were challenged as previously described (9) using vectors ± plasma, plasma, LPS (500 ng/ml), or IVIg® (Baxter SAS, Guyancourt, France). Plasma were pre-incubated individually with either HAdV-C5, D26 or B35 (at 1000 pp/cell) prior to *in vitro* MoDC challenge. When appropriate, plasma were classified into four groups according to their serological profile: (C5+, D26-), (C5+, D26-), (C5-, D26-) and (C5+, D26+) and pre-incubated with HAdV-C5 or D26 before the MoDC challenge. Plasma were tested repeatedly on MoDCs generated from different blood donors (n=3 to 6). Supernatants were collected after 14 h of culture and tested for TNF secretion by ELISA according to the manufacturer instructions (OptEIA human TNF ELISA Kit, BD Biosciences).

### Statistical analyses

All data were analysed using GraphPad Prism 5 software. Statistical analyses were determined using non-parametric Mann Whitney U test when two groups are compared and Kruskal Wallis ANOVA analyses for multiples inter-group comparisons. The non-parametric Spearman test correlation test was used to evaluate the association between NAb titers and TNF levels (r_s_ (rho): Spearman rank correlation). Correlation tests were performed with the online statistical tools for high-throughput data analysis (STHDA) (28).

## Results

### NAb titers for HAdV-C5, HAdV-D26, and HAdV-B35

We first evaluated the prevalence NAb in HIV-uninfected subjects from Burkina Faso, Chad, and France **(Figure 1 & 2B)**. Consistent with previous reports, the majority (60 ± 5%) of adults in all regions have HAdV-C5 NAbs, with median titers (IQR) of 643, 521 and 591 for Burkina Faso, Chad and France, respectively. For HAdV-D26, about 48% of individuals from the African countries have NAb, whereas found no sample with NAb titers ≥ 1000 in the French group. The median NAb titers were 407, 333 and 30 for Burkina Faso, Chad and France, respectively. By contrast, no sample showed significant titers for HAdV- B35 NAbs.

**Figure 1:**
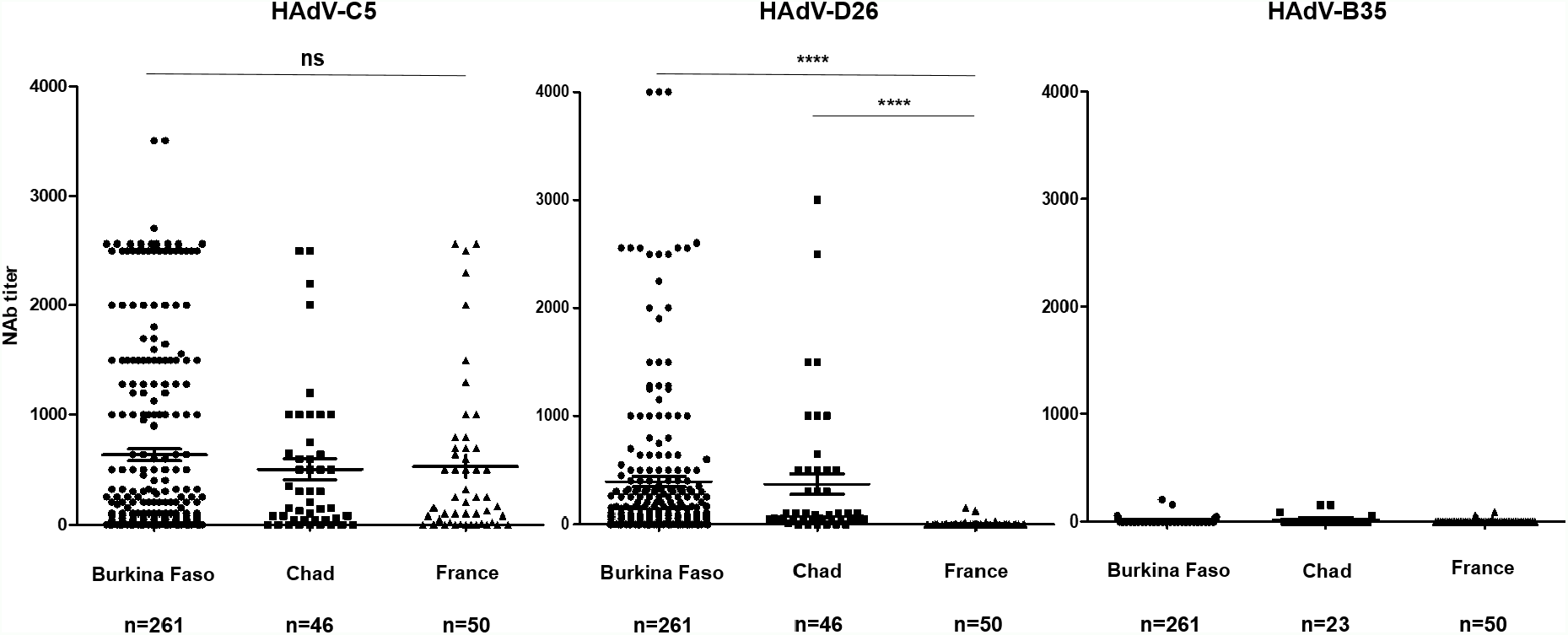
HAdV-C5, HAdV-D26, and HAdV-B35 NAb seroprevalence (bars: mean ± SEM) in healthy individuals from Burkina Faso, Chad and France. Statistical analyses were performed by Kruskal Wallis non-parametric Anova statistical test. p values are denoted as: ns: not significant, *p < 0.05; ** p < 0.01; *** p < 0.001 and **** p < 0.0001.

Individual NAb titers were stratified into three categories: ≤ 99 (negative or low), 100-999 (medium) and ≥ 1000 (high) **(Figures 2A & 2B)**. For HAdV-C5, 55%-66% of the African subjects had titers ≥ 100 and 22%-30% had Titers ≥ 1000. In France, ∼66% of the cohort had NAb titers ≥ 100 and ∼18% were ≥ 1000. For HAdV-D26, ∼48% of the individuals had NAb titers ≥ 100 in Burkina Faso and Chad, compared to only 4% in France. Additionally, ∼13% of the samples had high HAdV-D26 NAb titers in the sub-Saharan groups. Seroprevalence was as follows (NAb threshold ≥ 100): HAdV-C5 - 55% (143/261) and HAdV-D26 - 48% (126/261) in Burkina Faso; HAdV-C5 - 63% (29/46) and HAdV-D26 - 48% (22/46) in Chad, compared to HAdV-C5 - 66% (33/50) and HAdV-D26 4% (2/50) in the French cohort **(Figure 2B)**.

**Figure 2:**
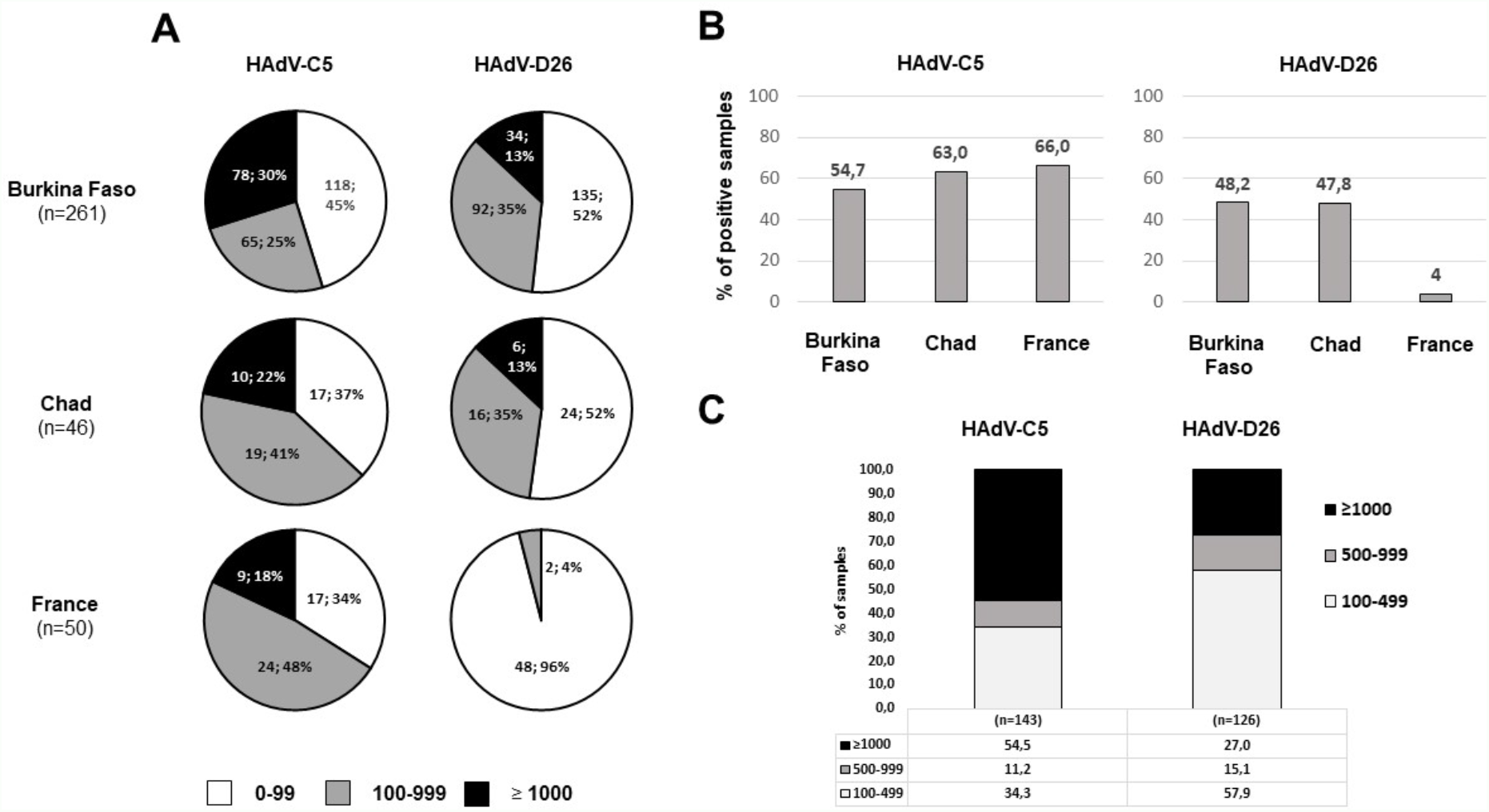
Distribution of HAdV-C5 and HAdV-D26 NAb titers in healthy individuals from Burkina Faso, Chad and France. **(A)** NAb titers were arbitrarily stratified into the following categories: ≤ 99, 100-999 and ≥ 1000. **(B)** Percentage of positive samples (NAb titer ≥ 100). **(C)** Distribution (percentage) of NAb titers among positive samples from Burkina Faso. NAb titer was divided into three groups: low (100-499), medium (500-999) and high (≥ 1000).

We found no samples with HAdV-B35 NAb titer greater than 100, and a seroprevalence < 6% for all groups tested **(data not shown). Figure 2C** shows the distribution of NAb titers among positive samples from Burkina Faso. NAb titres were stratified into three categories: low or moderate (100-499), medium (500-999) and high (≥ 1000). The majority of samples exhibited medium (11%) or high (54%) NAb titers for HAdV-C5 (65% total), while most of the cohort (58%) had low or moderate level of HAdV-D26 NAb. Nevertheless, 15% of the positive samples had medium NAb titers to HAdV-D26, and 27% had NAb titers ≥ 1000 (42% total).

### Correlation between NAb titers, sex, age, and HIV status in Burkina Faso

To better understand the natural history of HAdV infection among sub-Saharan populations, we explored possible relationships between HAdV seroprevalence and gender. We found no associations between single or dual-positive groups for HAdV-C5 and HAdV-D26 **(Figure S1A)**, and no differences in medians titer distribution according to sex (**Figure S1B & C)**. Assuming that HAdV type-specific NAb titers will wane over time, the comparison of seroprevalence between different age groups also does not indicate an age-dependent susceptibility to infections **(Figure S2A)** or the average NAb titers **(Figure S2B)**. However, seroprevalence for both HAdVs appears to increase overall with age, peaking in young adults and maintained throughout life.

### Immune-complexed (IC) HAdV-D26 activates MoDCs

We previously showed that IC-HAdV-C5 induces AIM2-associated pyroptosis in primary cultures of human phagocytes (29,30). To evaluate the impact of IC-HAdV-D26, we challenged MoDCs with plasma containing NAbs, preincubated with the respective HAdVs and then quantified TNF secretion. We divided plasma into groups according to their serological profile: single positive (HAdV-C5+/HAdV-D26-) and (HAdV-C5-/HAdV- D26+), dual positive (HAdV-C5+/HAdV-D26+) and dual negative (HAdV-C5-/HAdV-D26-). We also ensured that groups had similar sampling and comparable mean NAb titers. The HAdV-C5+/HAdV-D26- samples induced significantly higher production of TNF with HAdV-C5 compared to HAdV-D26, whereas the opposite was observed with the HAdV-C5- /HAdV-D26+ plasma group (**Figure 3A)**. The dual-positive group (HAdV-C5+/HAdV- D26+) induced significantly higher TNF production for both vectors in comparison to the dual-negative group (HAdV-C5-/HAdV-D26-). Additionally, TNF secretion for a given virus were comparable (regardless of the serological profile of the other virus).

**Figure 3:**
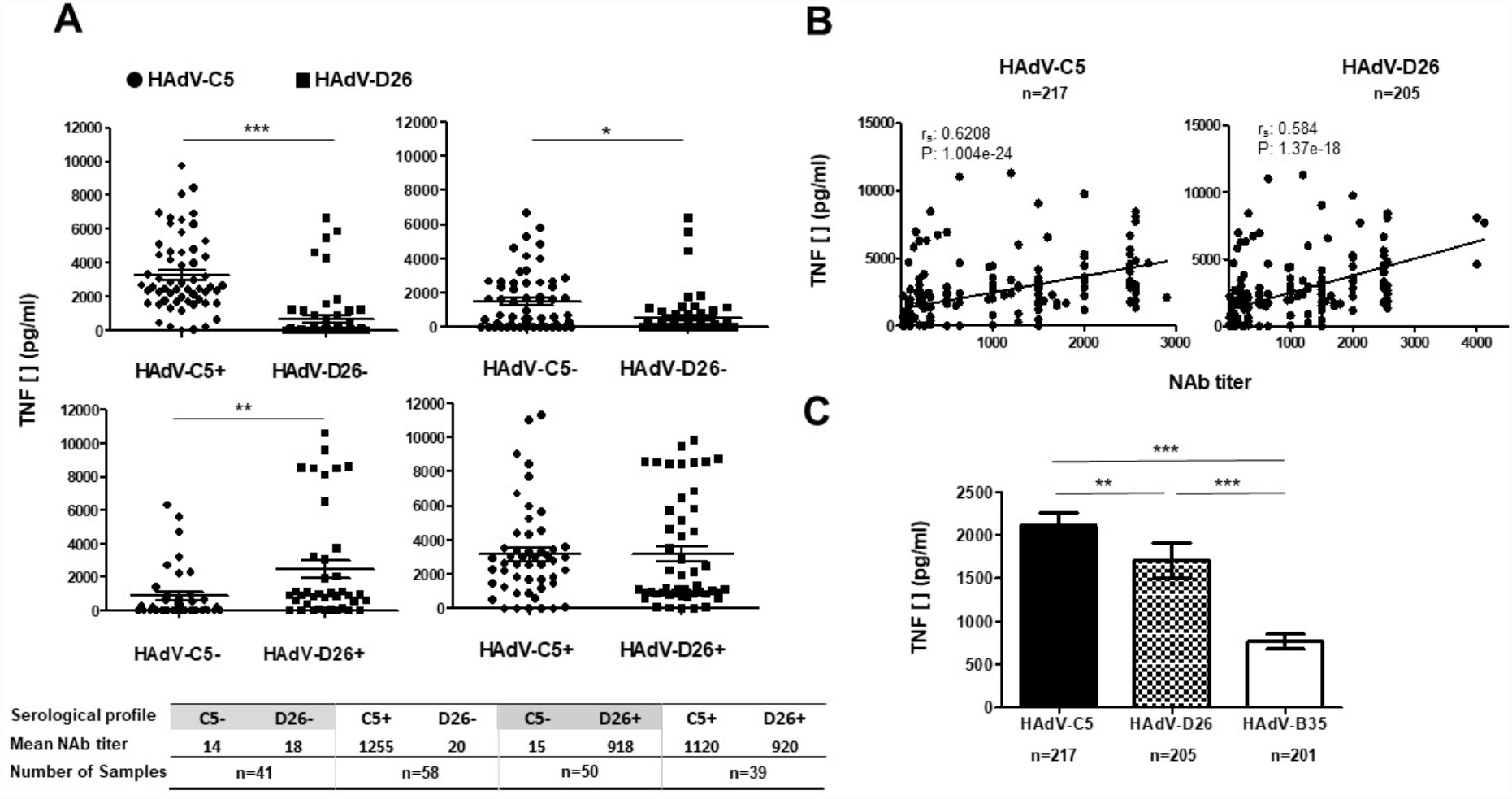
TNF secretion was used as a surrogate for MoDC activation following *in vitro* activation by various plasma from healthy individuals from Burkina Faso, pre-incubated with HAdV-C5, HAdV-D26 or HAdV-B35. **(A)** Individual and average (mean ± SEM) TNF secretion according to plasma serology profiles. Samples were classified into four groups: (C5+ D26+); (C5- D26-); (C5- D26+); (C5+ D26+) and individually pre-incubated with HAdV-C5 (•) or HAdV-D26 (◼). Positive samples (NAb titer ≥ 100). **(B)** Correlation between plasma NAb titers and TNF secretion by MoDC. **(C)** Mean (with SEM) TNF secretion by MoDC in presence of plasma individually pre-incubated with HAdV-C5, HAdV-D26 or HAdV-B35. Comparisons were performed by Kruskal Wallis non-parametric ANOVA statistical test. Correlation were performed with the non-parametric Spearman test. **r**_**s**_ (rho) = Spearman rank correlation. p values are denoted as: *p < 0.05; ** p < 0.01; and *** p < 0.001.

Monotonic regression analyses to examine the correlation between HAdVs NAb titers and the level of TNF secretion suggested modest correlation (r_2_: 0.62 and 0.58) between the IC-HAdV-C5 and -D26 **(Figure 3B)**. We then quantified the average production of TNF by MoDCs, pre-incubated with each plasma individually, and independently of their serological profile. This approach provides an overall estimate of the innate immune response to the HAdVs in a given population. The average TNF secretion level was significantly higher with HAdV-C5, followed by HAdV-D26 and HAdV-B35, respectively (**Figure 3C)**. Additionally, TNF secretion was significantly higher for HAdV-D26 compared with HAdV-B35. By contrast, MoDC activation with plasma from France (n=15) was modest with HAdV-D26 and HAdV-B35 compared to HAdV-C5 **(data not shown)**.

### HAdV seroprevalence and MoDC TNF secretion according to HIV status

HIV is a major burden in sub-Saharan Africa and HAdV infections are common in people with immunodeficiency, especially for HAdV-Ds, frequently isolated from immunocompromised individuals. To determine wether HAdV seroprevalence is linked to HIV status, we first evaluated the mean NAb titers in HIV-infected versus uninfected controls **(Figure 4A)**. For samples collected in Burkina Faso, mean NAb titers for HAdV- C5 only were significantly higher in the HIV-infected group. However, in Chad, this increase was observed for the three HAdV tested. Of note, seroprevalence reached ∼90% for HAdV-C5 and HAdV-D26 and 24% for HAdV-B35, compared to 63%, 48%, and 0% for uninfected controls **(Figure 2B & 4A)**. In addition, HIV-infected subjects tended to have a higher proportion of high NAb titers for both HAdV-C5 52% (25/46) and HAdV-D26 39% (18/46) **(data not shown)** compared to the HIV-uninfected groups **(Figure 2A)**.

**Figure 4:**
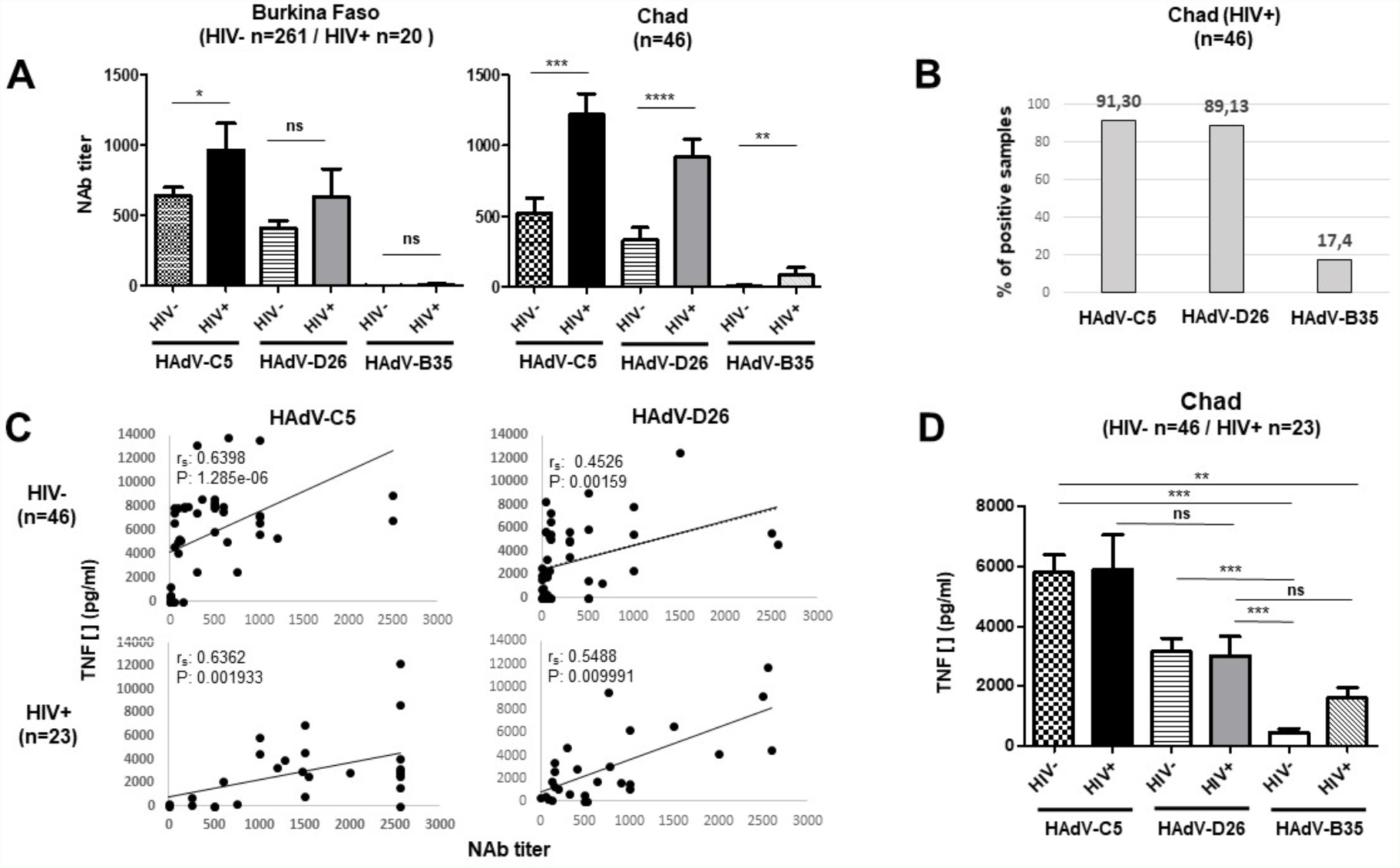
Overall HAdV-C5, HAdV-D26 and HAdV-B35 NAb titers and TNF secretion by MoDC challenged with plasma from HIV-infected (HIV+) individuals or uninfected controls (HIV-) from Burkina Faso or Chad, pre-incubated with various HAdVs. **(A)** Mean (with SEM) NAb titers among HIV-infected and HIV-uninfected populations. **(B)** Percentage of positive samples among HIV-infected subjects in Chad (NAb titer ≥ 100). **(C)** Correlation between plasma NAb titers and TNF secretion and **(D)** mean (with SEM) TNF secretion by MoDC challenged with plasma from healthy or HIV+ groups from Chad, pre-incubated with HAdV- C5, HAdV-D26 or HAdV-35. Statistical analysis were made using Mann Whitney or Kruskal Wallis ANOVA non-parametric tests for comparisons between two or multiples groups, respectively. Correlations were made with the non-parametric Spearman test. r_s_ (rho) = Spearman rank correlation. p values are denoted as: ns: not significant, *p < 0.05; ** p < 0.01; *** p < 0.001 and **** p < 0.0001.

Using IC-HAdVs (plasma + HAdVs) we then analysed the correlation between NAb titers and TNF production by MoDC for samples collected from people HIV-infected and uninfected individuals from Chad. In concordance with our previous observation **(Figure 3B)**, we found a positive but moderate correlation **(Figure 4C)** that was higher when plasmas where pre-incubated with HAdV-C5 compared to HAdV-D26. Finally, we compared the ability of plasmas to induce TNF secretion from MoDCs according to their HIV status. We found no difference in either of the HAdVs tested for stimulation between the HIV-infected and uninfected groups **(Figure 4D)**. However, we observed by comparison that plasma samples incubated with HAdV-C5 or HAdV-D26 induced greater TNF levels compared to HAdV-B35, regardless of the HIV status. No significant differences were found between HAdV-C5 and HAdV-D26. For HAdV-B35, only the HIV-uninfected group reached a significant variation compared with HAdV-D26. Finally, we observed that TNF production was ∼3-fold higher in samples from Chad compared to Burkina Faso, regardless of HIV serologic profile and for all three HAdVs tested **(Figure 3C & 4D)**.

## Discussion

All vaccines, and in particular those derived from HAdVs, must be vetted judiciously (31). We are just beginning to understand the biological mechanisms that govern the interactions between pre-existing immunity, alarmins, coagulation factors and the effectiveness of HAdV vectors (32). In addition to affecting the efficacy of the vaccine, the adverse events of the STEP/Phambili trials from the HIV Vaccine Trials Network (HVTN) illustrate the need to take precautions (33). In this study, we found high HAdV-C5 seroprevalences as previously reported in other sub-Saharan regions (11,20–22). However, for HAdV-D26, limited information was available. Our results on the French group confirmed a low HAdV- D26 seroprevalence in Europe (21). Investigations performed on rural populations from other sub-Saharan African countries showed relative high prevalence and genetic diversity among the species D of HAdV (HAdV-Ds) (34). This study also suggested that the fecal-oral transmission, hygiene measures, nutrition and local infectious burden play a role in HAdV-Ds spread.

The majority of studies on HAdV-D26 or -B35 seroprevalence were conducted by pharmaceutical companies to monitor the clinical applicability and of HAdV in a context of vaccine development. We found a higher HAd-C5 and -D26 seroprevalence and mean NAb titers in sub-Saharan cohorts compared to France (20–22). One study, however, found very high NAb seroprevalence and titers for HAdV-D26 in cohorts from Cameroun, similar to or higher than HAdV-C5 (19). These data also highlight the discrepancy between studies regarding the level of HAdV-D26 seroprevalence in African cohorts. Taken together, these studies showed that HAdV-D26 seroprevalence in sub-Saharan zone ranges from 45-88%. However, the studies also generally showed that the proportion of people with a high NAb titer (≥1000) for HAdV-D26 was low (3.4% overall with a maximum of 6.3% in South Africa) (21,22,35). Within our groups, we found a relatively high proportion of individuals with HAdV-D26 NAb titers above those anticipated affecting the performance of a HAdV-based vaccines **(Figure 2C)**. The proportion of samples having NAb titers ≥ 1000 was the highest in the HIV-infected group from Chad, reaching ∼40%.

HAdV-B35 seroprevalence was low for all groups tested **(Figures 1A & 2A)**. Consistent with other studies, seroprevalence for all HAdVs was independent of the donor’s gender (36). For the pediatric population, outcomes are sometimes contradictory, indicating either lower seroprevalence compared to adults (36), or that infections/seroconversion occurred relatively early in life, then waned with age (21,37). In our study, we found that NAb serprevalence increased with age, but the mean NAb titer did not vary significantly **(Figures S2A & S2B)**.

HIV is a major burden in sub-Saharan Africa. In addition, HAdV infections are common in people with immunodeficiency (38) and can cause significant morbidity, especially for HAdV-Ds (39). Asymptomatic HAdV infections are also frequent in HIV-infected patients who are not on antiretroviral therapy. However, most studies conducted on HAdV-C5 and HIV status showed a comparable seroprevalence between HIV-infected and non-infected individuals, which remains constant over the course of the progression to AIDS (40). For HAdV-D26 or HAdV-B35, the correlation between HIV status and seroprevalence was rarely described. Older studies showed that HAdV-B35 was preferentially isolated from immunocompromised patients with AIDS (41), but did not indicate significant variations in subjects from the United States or Africa, for both HAdV-D26 and HAdV-B35 (22,42). In China, however, HAdV-D26 seroprevalence is higher in the population at high risk for HIV infection than in the low risk cohort (43). In our study, HAdV-C5 seroprevalence was significantly higher for the HIV-infected groups compared to the general population in Burkina Faso and for all three HAdVs tested in Chad **(Figure 4A)**. Despite numerous reports reflecting the importance of HAdV in patients with AIDS, the opportunistic role of this pathogen is poorly documented and, considering these data, may be underestimated in sub-Saharan Africa.

To better understand the consequences of HAdV NAbs, we used an *ex vivo* model based on MoDC activation (9). With this system, we show consistent TNF secretion according to the HAdV serological profiles **(Figure 3A)**. However, some samples occasionally induced MoDC activation even though they were HAdV-NAb negative/low. This observation was confirmed by the moderate correlation observed between the NAb titers of the samples and the level of MoDC activation **(Figure 3B)**. We hypothesize that non-NAbs can form complexes and activate MoDC. It has been established that HAdV humoral response is directed against multiple capsid components (44). The dogma in the field suggests that NAbs induced by HAdV natural infection lead to poor cross-reactivity, but this has not been systematically analysed. In addition, studies have shown a probable cross-protection between HAdVs from the same species (45,46).

We previously found that the amount of TNF secreted reflects the activation of the host immune response and could therefore be indicative of vaccine efficacy. The Inflammatory response induced by HAdV-D26 and HAdV-B35 are generally lower in comparison to HAdV-C5, which correlates with the seroprevalence profiles observed for these populations **(Figure 3C)**. However, the differences between HAdVs were less evident for samples collected in Chad **(Figure 4D)**. Our analyses showed similar TNF production for all three HAdVs in HIV-infected participants. The amount of TNF secreted by MoDC was also higher in subjects from Chad compared to Burkina Faso in general. We presume the possibility of numerous cross-reactions, or the involvement of other factors linked to environment and exposure to infectious agents, but also to genetic variations, which may influence the immune response to HAdVs-derived vaccines (47). In addition, several extracellular proteins in plasma interact with HAdV and can increase the uptake by and activation of DC (48).

Will HAdV-D26 or -B35-based vaccines prime individuals for infection by HIV? (49). According to the vaccine manufacturers, there is no clinical evidence to date that pre-existing immunity, whether natural or vector-induced, influences or blunts the efficacy of HAdV-D26 or B35-derived vaccines (12). These studies suggested that the major difference with HAdV-C5 could be explained by biological variables such as receptor usage (12). However, we demonstrate that individuals with high HAdV-D26 NAb titers are common in some sub-Saharan areas, with rates comparable to those of HAdV-C5. We found that plasma from people HIV-infected + HAdV-C5 or HAdV-D26 can efficiently activate MoDC. Thus, we share the concerns about the use of HAdV-based vaccines in sub-Saharan zones with high HIV endemic populations (49,50). The risk/benefit assessment of HAdV-derived vaccines must address HAdVs seroprevalence. We also recommend surveillance of de novo HIV infections in high HIV-prevalent populations. In addition, our study underscores the need to pre-evaluate the efficacy and safety of vaccines in sub-Saharan populations. These recommendations are all the more important as we may be living through a crucial and unique turning point in the history of vaccination, which could influence future strategies to reduce vaccine hesitancy worldwide.

## Data Availability

All data produced in the present study are available upon reasonable request to the authors.
All data produced in the present work are contained in the manuscript.

## Acknowledgments

We thank Mehdi Taleb, Sofia Salazar Arenas, Chloé Houques, Lumen Ishimwe, Aboudou Djobo, Lucas August, Marina Lavigne, Hiep Tran, Coraline Cheneau, TT Phuong Tran, Karsten Eichholz, Karine Bollore et Jean Pierre Molès for reagents and technical help. We thank Thierry Gostan and Myriam Boyer for their help on statistical analyses and flow cytometry, respectively. EJK is an INSERM fellow. We are grateful to Eric A. Weaver and Andre Lieber for the gift of HAdV-D26 and HAdV-B35, respectively.

## Biographical Sketch of the first author

Dr Ouoba is a pharmacist and obtained a PhD in immunology/virology from the University of Montpellier, France. She works on diseases with epidemic potential at the Muraz research centre in Bobo Doulasso, Burkina Faso. Her main research interests are the detection and prevention of viral infections in sub-Saharan areas.

## Supplementary data

**Figure S1:**
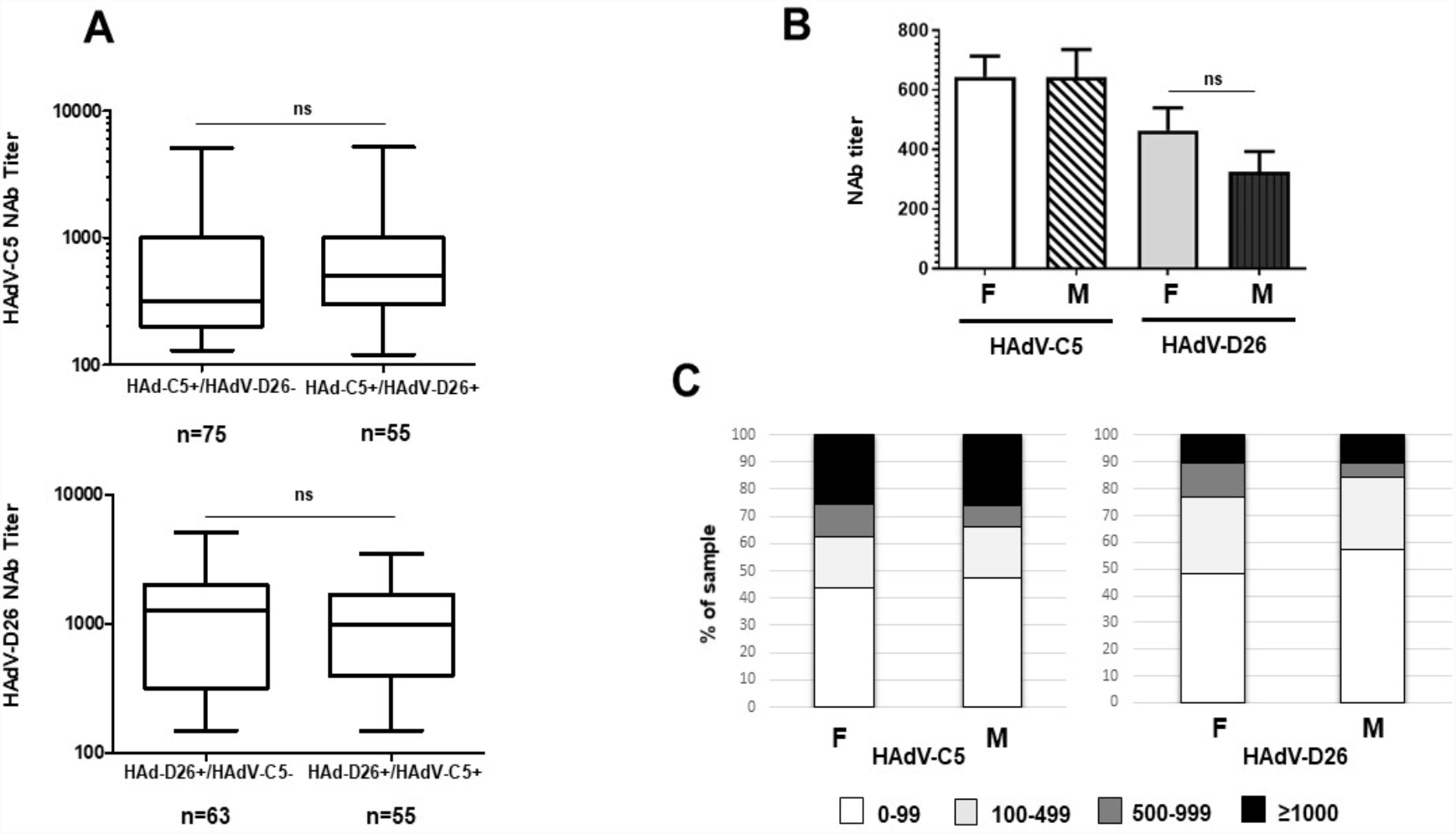
Overall HAdV-C5 and HAdV-D26 NAb titers among healthy individuals from Burkina Faso according to **(A)** single- or dual-positive groups (box-plots shows the minimum, first quartile, median, third quartile and maximum titer levels) and **(B)** gender groups (mean with SEM). **(C)** Distribution of NAb titers among males (n = 107) and females (n = 154) groups. NAb titers was stratified into 4 groups: negative (0-99), low (100-499), medium (500- 999) and high (≥ 1000). Comparison was performed using the non-parametric Mann Whitney statistical test. ns: not significant, *p < 0.05; ** p < 0.01; *** p < 0.001.

**Figure S2:**
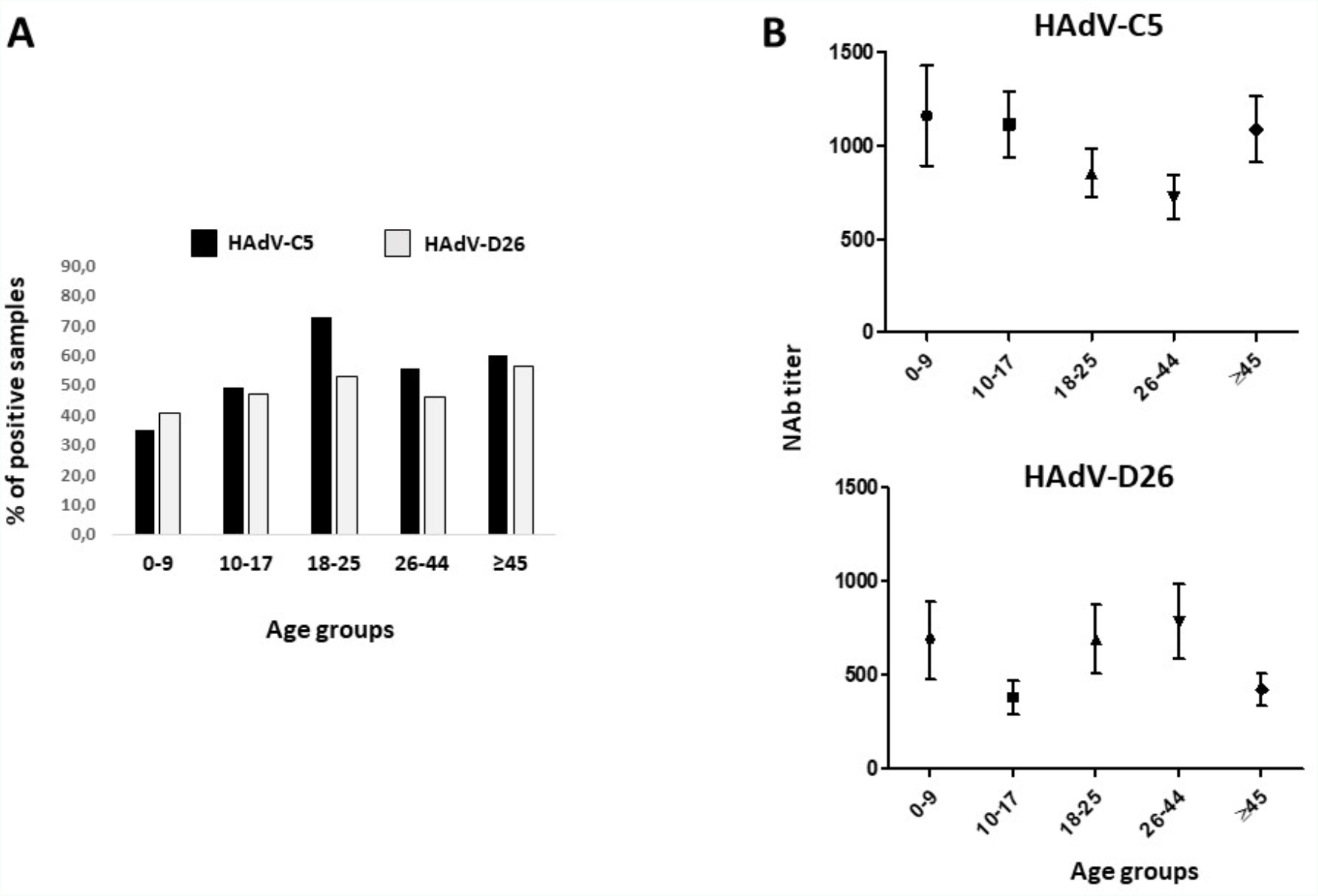
NAb seroprevalence against HAdV-C5 and HAdV-D26 among different age groups of healthy individuals from Burkina Faso. **(A)** Percentage of positive samples (NAb titer ≥ 100) among different age groups for HAdV-C5 (Black bar) and HAdV-D26 (grey bar). **(B)** Mean NAb seroprevalence (mean ± range) among different age groups of healthy positive subjects from Burkina Faso. Comparisons were performed by Kruskal Wallis non-parametric ANOVA statistical test.

